# Strong Association between Infectious Mononucleosis and Cardiac Allograft Vasculopathy Studying Risk Factors and Viral Infections

**DOI:** 10.1101/2025.02.18.25322503

**Authors:** Mohammad Reza Movahed, Mohammad Javad Namazi, Mahsa Rezasoltani, Mehrtash Hashemzadeh

## Abstract

**Background:** Cardiac Allograft vasculopathy (CAV) is a significant cause of late transplant failure. Using a large database, the study’s objective was to assess traditional and infectious risk factors linked to the occurrence and severity of CAV.

**Method:** Using the large inpatient sample database (NIS), we evaluated any association between CAV and risk factors and infectious viral agents. Additionally, we assessed the severity of CAV based on the occurrence of revascularizations.

**Results:** A total of 78,330 heart transplant recipients were identified. CAV was diagnosed in 1,015 patients overall. Patients with CAV had a higher mortality rate (4.4% vs 2.1%, OR: 2.09 CI 1.08-4.03 p=0.03). All known traditional risk factors and baseline characteristics, including gender, race, hypertension, hyperlipidemia, diabetes mellitus, and smoking, were not linked to the existence of CAV, except for being younger (mean age 56 vs 59 years). Furthermore, a history of infectious mononucleosis strongly correlated with CAV (OR:8.9 CI 2.68–29.6 p<0.001). Being younger not only increases the possibility of the development of CAV but also increases the probability of undergoing coronary bypass surgery after a heart transplant. Influenza and other forms of viral infections, such as Cytomegalovirus, did not correlate with the presence of CAV.

**Conclusion:** Younger age was associated with CAV but no other traditional risk factors. Infectious mononucleosis, the only infectious agent correlating with CAV, had a very high association with CAV, warranting further investigation.

## Introduction

Heart transplant recipients should be very concerned about cardiac allograft vasculopathy (CAV) since it can seriously compromise long-term survival and increase the likelihood of transplant failure. CAV is characterized as new atherosclerosis development in a transplanted heart that can lead to ischemia and even graft failure.

CAV continues to be the primary cause of death after a heart transplant, even with advancements in immunosuppressive therapy ^(1)^. It is essential to understand the variables linked to the occurrence and intensity of this illness to improve patient outcomes and create focused therapies. This study aims to evaluate any correlation between the diagnosis of CAV and traditional risk factors and viral infections. Furthermore, the severity was assessed based on the occurrence of percutaneous coronary intervention (PCI) and coronary bypass surgery (CABG).

## Methods

For our analysis, we used the Nationwide Inpatient Sample (NIS) database, which is the largest all-payer inpatient healthcare database in the United States. Comprehensive information about hospital inpatient stays is available in the NIS database, facilitating the analysis of different illnesses and treatments. Using NIS, we evaluated any association between the presence of AGV and traditional cardiovascular risk factors and baseline characteristics in adult inpatients over age 18. Additionally, we assessed the severity of CAV based on the occurrence of PCI or CABG. Traditional risk factors are age, gender, race, hypertension, hyperlipidemia, diabetes mellitus, and smoking status. We used the ICD-10 code of T86.290 for cardiac allograft vasculopathy.

### Statistical analysis

For continuous data, the mean and standard deviation were used to describe patient demographic, clinical, and hospital characteristics; for categorical variables, proportions with 95% confidence intervals were used. To compare continuous variables, the two-sample t-test will be employed, and to compare categorical variables, the Chi-squared/Fisher’s Exact Test will be utilized. We used logistic regression to determine the odds of binary clinical outcomes due to patient and hospital factors. The probability of the result (Odds Ratios (95% CI)) arising between the characteristics was determined using multiple logistic regression. After applying population discharge weights, analyses were conducted. All p-values were calculated as 2-sided, with a significance level set at p-value< 0.05. The software STATA17 (Stata Corporation, College Station, TX) was used for data analysis.

## Results

CAV was found in 1,015 (1%) of the 78,330 heart transplant patients. We found no significant correlation between the presence of CAV and traditional risk factors, such as gender, race, hypertension, hyperlipidemia, diabetes mellitus, and smoking, except for being younger (mean age 56 vs 59 years). Furthermore, infectious mononucleosis was highly and significantly more than 8 times associated with the occurrence of CAV (OR: 8.91 CI: 2.68-29.60, p<0.001). No correlation was found between CAV with any other infectious agents. (Table 1, Figure 1).

**Figure 1:**
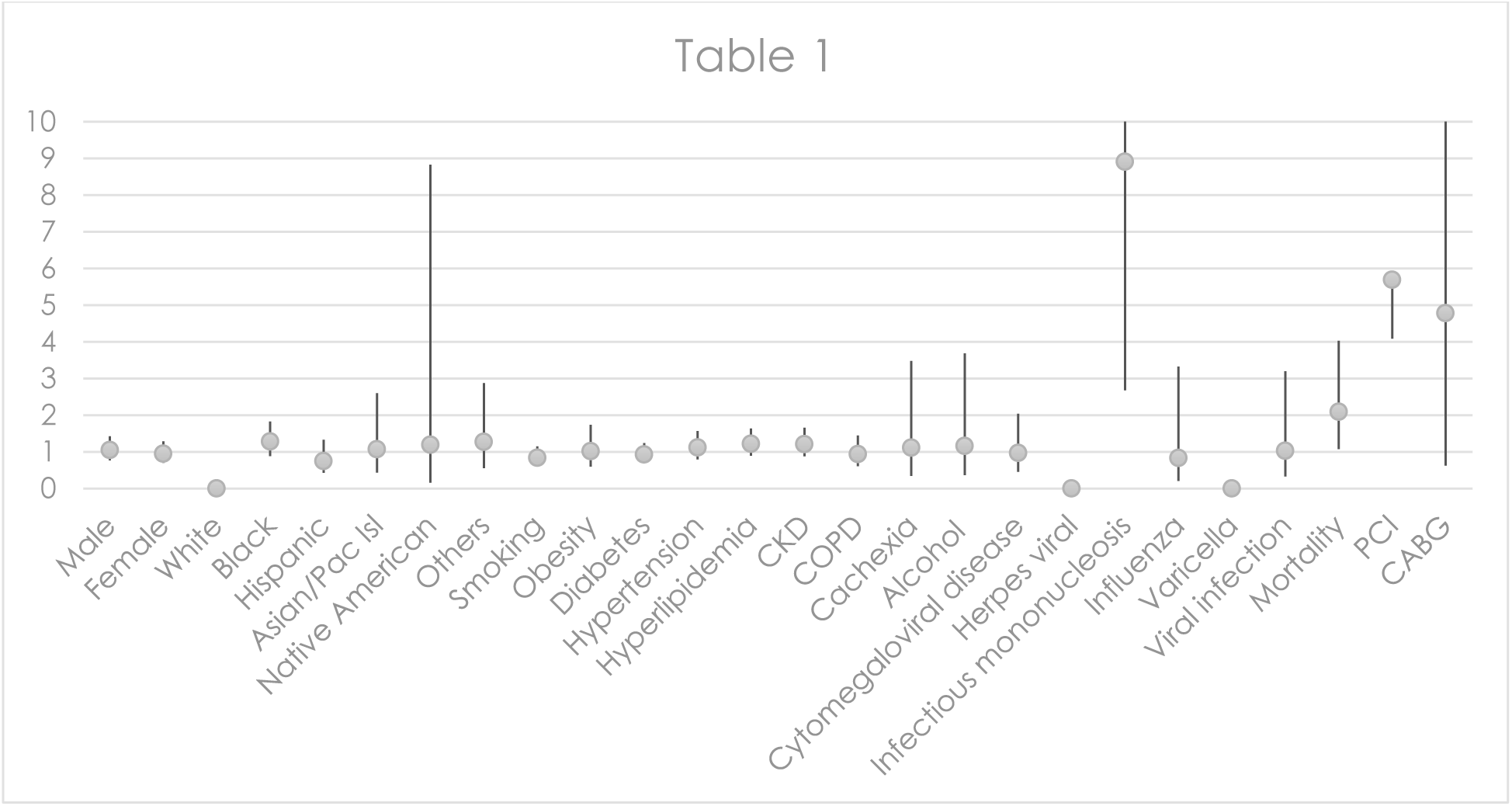
Odds ratios for associations between cardiac allograft vasculopathy (CAV) with infectious agents and clinical characteristics

**Table 1:**
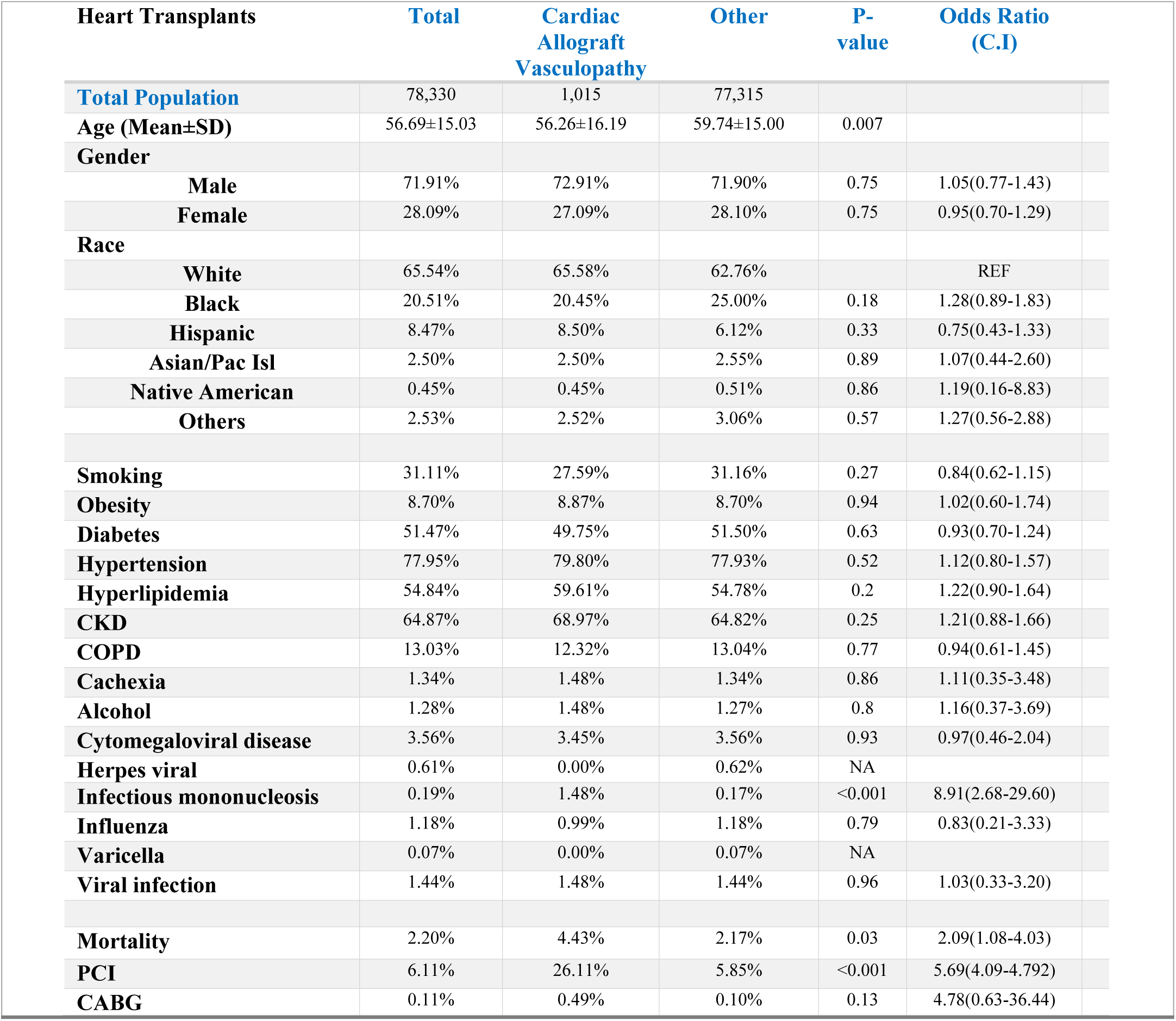
Baseline characteristics of patients with and without CAV in heart transplant patients.

| Heart Transplants | Total | Cardiac<br>Allograft<br>Vasculopathy | Other | P-value | Odds Ratio<br>(C.I) |
| --- | --- | --- | --- | --- | --- |
| **Total Population** | 78,330 | 1,015 | 77,315 |  |  |
| **Age (Mean±SD)** | 56.69±15.03 | 56.26±16.19 | 59.74±15.00 | 0.007 |  |
| **Gender** |  |  |  |  |  |
| **Male** | 71.91% | 72.91% | 71.90% | 0.75 | 1.05(0.77-1.43) |
| **Female** | 28.09% | 27.09% | 28.10% | 0.75 | 0.95(0.70-1.29) |
| **Race** |  |  |  |  |  |
| **White** | 65.54% | 65.58% | 62.76% |  | REF |
| **Black** | 20.51% | 20.45% | 25.00% | 0.18 | 1.28(0.89-1.83) |
| **Hispanic** | 8.47% | 8.50% | 6.12% | 0.33 | 0.75(0.43-1.33) |
| **Asian/Pac Isl** | 2.50% | 2.50% | 2.55% | 0.89 | 1.07(0.44-2.60) |
| **Native American** | 0.45% | 0.45% | 0.51% | 0.86 | 1.19(0.16-8.83) |
| **Others** | 2.53% | 2.52% | 3.06% | 0.57 | 1.27(0.56-2.88) |
| **Smoking** | 31.11% | 27.59% | 31.16% | 0.27 | 0.84(0.62-1.15) |
| **Obesity** | 8.70% | 8.87% | 8.70% | 0.94 | 1.02(0.60-1.74) |
| **Diabetes** | 51.47% | 49.75% | 51.50% | 0.63 | 0.93(0.70-1.24) |
| **Hypertension** | 77.95% | 79.80% | 77.93% | 0.52 | 1.12(0.80-1.57) |
| **Hyperlipidemia** | 54.84% | 59.61% | 54.78% | 0.2 | 1.22(0.90-1.64) |
| **CKD** | 64.87% | 68.97% | 64.82% | 0.25 | 1.21(0.88-1.66) |
| **COPD** | 13.03% | 12.32% | 13.04% | 0.77 | 0.94(0.61-1.45) |
| **Cachexia** | 1.34% | 1.48% | 1.34% | 0.86 | 1.11(0.35-3.48) |
| **Alcohol** | 1.28% | 1.48% | 1.27% | 0.8 | 1.16(0.37-3.69) |
| **Cytomegaloviral disease** | 3.56% | 3.45% | 3.56% | 0.93 | 0.97(0.46-2.04) |
| **Herpes viral** | 0.61% | 0.00% | 0.62% | NA |  |
| **Infectious mononucleosis** | 0.19% | 1.48% | 0.17% | <0.001 | 8.91(2.68-29.60) |
| **Influenza** | 1.18% | 0.99% | 1.18% | 0.79 | 0.83(0.21-3.33) |
| **Varicella** | 0.07% | 0.00% | 0.07% | NA |  |
| **Viral infection** | 1.44% | 1.48% | 1.44% | 0.96 | 1.03(0.33-3.20) |
| **Mortality** | 2.20% | 4.43% | 2.17% | 0.03 | 2.09(1.08-4.03) |
| **PCI** | 6.11% | 26.11% | 5.85% | <0.001 | 5.69(4.09-4.792) |
| **CABG** | 0.11% | 0.49% | 0.10% | 0.13 | 4.78(0.63-36.44) |

When analyzing the severity of CAV, certain traditional risk variables become relevant, as shown by the need for PCI and CABG. There is an independent correlation between smoking and a higher chance of requiring PCI in patients with CAV. Smoking (OR: 2.16, CI: 1.11-4.18, p-value=0.02, table 2). Patients with CAV had a death rate that was twice as high as those without it (4.4% vs. 2.1%, OR: 2.09, CI: 1.08–4.03, p = 0.03). Mortality was higher in older patients with CAV (mean age of death was 69.89±7.14 vs 55.62±16.26 who survived, p < 0.001, table 3). Furthermore, younger recipients were more likely to have CAV (age, p=0.007) (Table 1) and more likely to undergo CABG (Table 4).

**Table 2:**
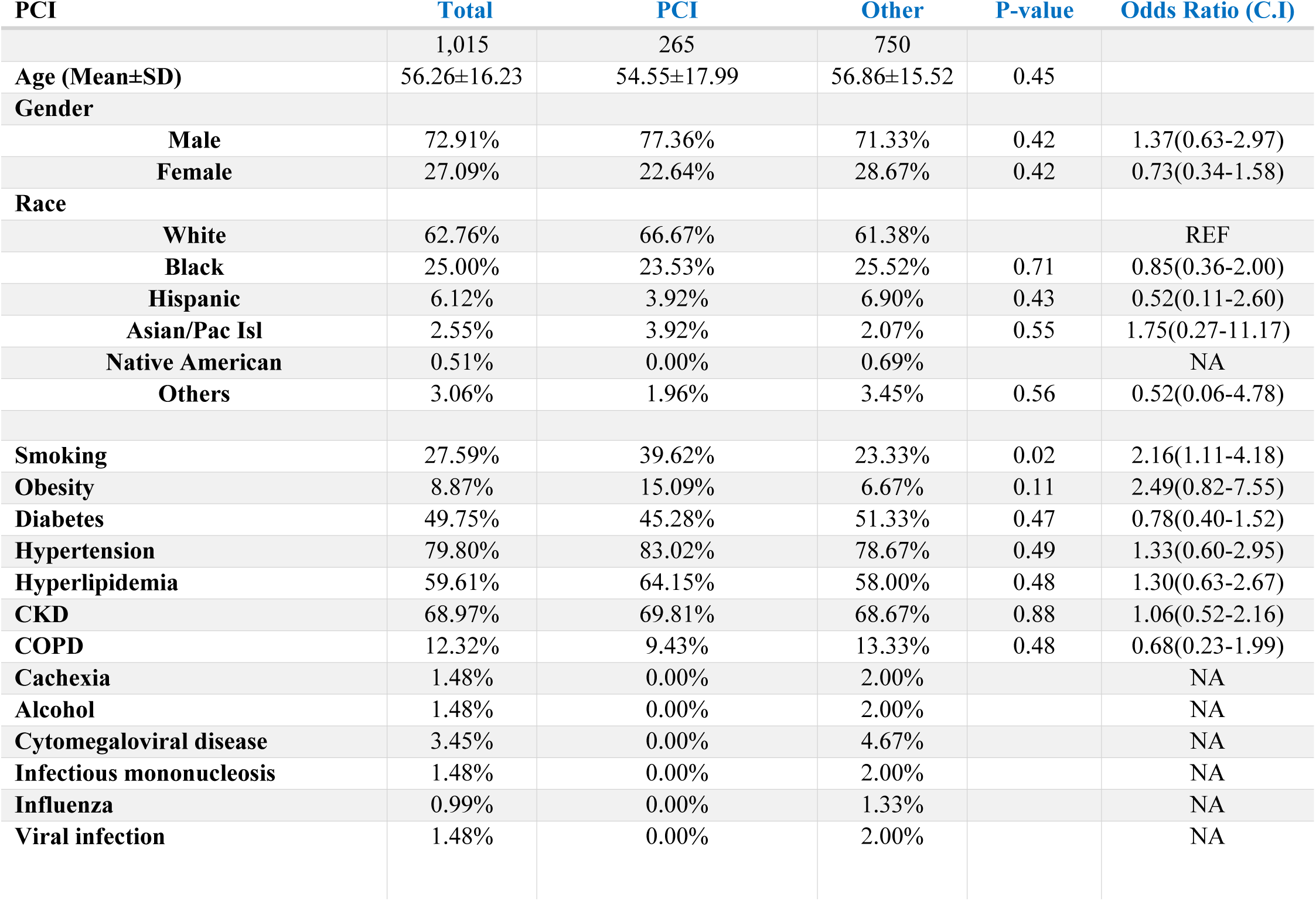
The likelihood of performing percutaneous coronary intervention (PCI) in heart transplant patients with cardiac allograft vasculopathy (CAV).

| PCI | Total | PCI | Other | P-value | Odds Ratio (C.I) |
| --- | --- | --- | --- | --- | --- |
|  | 1,015 | 265 | 750 |  |  |
| Age (Mean±SD) | 56.26±16.23 | 54.55±17.99 | 56.86±15.52 | 0.45 |  |
| Gender |  |  |  |  |  |
| Male | 72.91% | 77.36% | 71.33% | 0.42 | 1.37(0.63-2.97) |
| Female | 27.09% | 22.64% | 28.67% | 0.42 | 0.73(0.34-1.58) |
| Race |  |  |  |  |  |
| White | 62.76% | 66.67% | 61.38% |  | REF |
| Black | 25.00% | 23.53% | 25.52% | 0.71 | 0.85(0.36-2.00) |
| Hispanic | 6.12% | 3.92% | 6.90% | 0.43 | 0.52(0.11-2.60) |
| Asian/Pac Isl | 2.55% | 3.92% | 2.07% | 0.55 | 1.75(0.27-11.17) |
| Native American | 0.51% | 0.00% | 0.69% |  | NA |
| Others | 3.06% | 1.96% | 3.45% | 0.56 | 0.52(0.06-4.78) |
| Smoking | 27.59% | 39.62% | 23.33% | 0.02 | 2.16(1.11-4.18) |
| Obesity | 8.87% | 15.09% | 6.67% | 0.11 | 2.49(0.82-7.55) |
| Diabetes | 49.75% | 45.28% | 51.33% | 0.47 | 0.78(0.40-1.52) |
| Hypertension | 79.80% | 83.02% | 78.67% | 0.49 | 1.33(0.60-2.95) |
| Hyperlipidemia | 59.61% | 64.15% | 58.00% | 0.48 | 1.30(0.63-2.67) |
| CKD | 68.97% | 69.81% | 68.67% | 0.88 | 1.06(0.52-2.16) |
| COPD | 12.32% | 9.43% | 13.33% | 0.48 | 0.68(0.23-1.99) |
| Cachexia | 1.48% | 0.00% | 2.00% |  | NA |
| Alcohol | 1.48% | 0.00% | 2.00% |  | NA |
| Cytomegaloviral disease | 3.45% | 0.00% | 4.67% |  | NA |
| Infectious mononucleosis | 1.48% | 0.00% | 2.00% |  | NA |
| Influenza | 0.99% | 0.00% | 1.33% |  | NA |
| Viral infection | 1.48% | 0.00% | 2.00% |  | NA |

**Table 3:** Mortality of CAV based in demographics.

| **Mortality** | **Total** | **DIED** | **Other** | **P-value** | **Odds Ratio (C.I)** |
| --- | --- | --- | --- | --- | --- |
| **Total Population** | 1,015 | 45 | 970 |  |  |
| **Age (Mean±SD)** | 56.26±16.23 | 69.89±7.14 | 55.62±16.26 | <0.001 |  |
| **Gender** |  |  |  |  |  |
| **Male** | 72.91% | 55.56% | 73.71% | 0.24 | 0.45(0.12-1.72) |
| **Female** | 27.09% | 44.44% | 26.29% | 0.24 | 2.24(0.58-8.64) |
| **Race** |  |  |  |  |  |
| **White** | 62.76% | 55.56% | 63.10% |  | REF |
| **Black** | 25.00% | 22.22% | 25.13% | 1 | 1.00(0.18-5.62) |
| **Hispanic** | 6.12% | 11.11% | 5.88% | 0.5 | 2.15(0.23-20.32) |
| **Asian/Pac Isl** | 2.55% | 0.00% | 2.67% |  | NA |
| **Native American** | 0.51% | 0.00% | 0.53% |  | NA |
| **Others** | 3.06% | 11.11% | 2.67% | 0.19 | 4.72(0.45-49.47) |
| **Smoking** | 27.59% | 0.00% | 28.87% |  | NA |
| **Obesity** | 8.87% | 11.11% | 8.76% | 0.81 | 1.30(0.15-11.35) |
| **Diabetes** | 49.75% | 44.44% | 50.00% | 0.74 | 0.80(0.21-3.03) |
| **Hypertension** | 79.80% | 66.67% | 80.41% | 0.33 | 0.49(0.11-2.09) |
| **Hyperlipidemia** | 59.61% | 44.44% | 60.31% | 0.34 | 0.53(0.14-2.00) |
| **CKD** | 68.97% | 100.00% | 67.53% |  | NA |
| **COPD** | 12.32% | 22.22% | 11.86% | 0.37 | 2.12(0.41-10.93) |
| **Cachexia** | 1.48% | 0.00% | 1.55% |  | NA |
| **Alcohol** | 1.48% | 0.00% | 1.55% |  | NA |
| **Cytomegaloviral disease** | 3.45% | 11.11% | 3.09% | 0.23 | 3.92(0.42-36.95) |
| **Infectious mononucleosis** | 1.48% | 0.00% | 1.55% |  | NA |
| **Influenza** | 0.99% | 0.00% | 1.03% |  | NA |
| **Viral infection** | 1.48% | 0.00% | 1.55% |  | NA |

**Table 4:** Association of Coronary bypass grafting (CABG) performed for patients with CAV based on demographics and risk factors.

| **CABG** | **Total** | **CABG** | **Other** |
| --- | --- | --- | --- |
| **Total Population** | 1,015 | 5 | 1,010 |
| **Age (Mean±SD)** | 56.26±16.23 | 52.00±0 | 56.28±16.27 |
| **Gender** |  |  |  |
| **Male** | 72.91% | 100.00% | 72.77% |
| **Female** | 27.09% | 0.00% | 27.23% |
| **Race** |  |  |  |
| **White** | 62.76% | 100.00% | 62.56% |
| **Black** | 25.00% | 0.00% | 25.13% |
| **Hispanic** | 6.12% | 0.00% | 6.15% |
| **Asian/Pac Isl** | 2.55% | 0.00% | 2.56% |
| **Native American** | 0.51% | 0.00% | 0.51% |
| **Others** | 3.06% | 0.00% | 3.08% |
| **Smoking** | 27.59% | 100.00% | 27.23% |
| **Obesity** | 8.87% | 0.00% | 8.91% |
| **Diabetes** | 49.75% | 100.00% | 49.50% |
| **Hypertension** | 79.80% | 100.00% | 79.70% |
| **Hyperlipidemia** | 59.61% | 100.00% | 59.41% |
| **CKD** | 68.97% | 100.00% | 68.81% |
| **COPD** | 12.32% | 0.00% | 12.38% |
| **Cachexia** | 1.48% | 0.00% | 1.49% |
| **Alcohol** | 1.48% | 0.00% | 1.49% |
| **Cytomegaloviral disease** | 3.45% | 0.00% | 3.47% |
| **Infectious mononucleosis** | 1.48% | 0.00% | 1.49% |
| **Influenza** | 0.99% | 0.00% | 0.99% |
| **Viral infection** | 1.48% | 0.00% | 1.49% |

## Discussion

Patients with CAV after heart transplantation are at risk for an unusual, accelerated form of coronary vascular disease. This complication is a significant contributor to medium- to long-term morbidity and mortality following transplantation. One in three patients had CAV five years after the transplant, demonstrating an elevated incidence of CAV. Although several immunological and non-immune variables have been identified, it is still unknown what the precise pathogenic mechanisms, linkages, and relative importance of these variables are in the development of CAV. ^(2)^ Thirty to fifty percent of patients experience CAV five years following heart transplantation ^(3)^. Treatment consists of PCI, CABG, and heart re-transplantation ^(4)^. Re-transplantation is one of the significant treatments now available for CAV, but its use is limited because there are no suitable matches for organ donors. Individuals whose CAV worsens after revascularization may benefit from re-transplantation ^(5)^. Some studies have looked into the possible risk factors that allograft patients might have for CABG and PCI. ^(6)^

Factors such as immunological response, compatibility between the donor and recipient, and the effectiveness of immunosuppressive medications may be more crucial in the development of CAV. ^(7)^

Our study validates traditional risk factors for allograft vasculopathy, which include smoking, hypertension, and hyperlipidemia, for coronary artery disease in the general population ^(8,9)^.

There are several critical distinctions between atherosclerosis and CAV. The most important one is that atheroma’s intima typically lacks lipid-rich necrotic core and foam cells, but atheroma’s does. If observed, these might be pre-existing atheromas that developed in the donor before transplantation. Atheroma’s smooth muscle cells (SMCs) are grouped in a fibrous cap next to the endothelium cell (EC) lining. As opposed to the CAV arrangement, which localizes SMCs to the deeper sections of the intima close to the media and concentrates the inflammatory infiltrate subjacent to the luminal endothelium, T cells, and macrophages are primarily localized to shoulder regions of the lesion. These similarities and differences are readily explained if one posits that adaptive immune responses to antigens drive both processes but that the antigens are different. ^(10)^.

In contrast, the principal antigens in the case of CAV are non-self-major histocompatibility complex molecules, especially HLA-DR, expressed most abundantly on the luminal EC ^(11)^. This hypothesis explains the distinct localization sites of the inflammatory infiltrates. It explains why atherosclerosis is a systemic disease and why CAV stops at the suture lines separating the donor from the host ^(12)^. With these existing differences, this is expected as our data demonstrates most traditional risk factors such as diabetes(p-value= 0.63), hypertension(p-value=0.52), hyperlipidemia(p-value=0.2), obesity(p-value=0.94), alcohol (p-value=0.8) same as CKD(p-value= 0.25, p-value= 0.88), COPD(p-value=0.77, p-value= 0.48), Cachexia (p-value= 0.86) did not correlate to development and progression of CAV respectively or have small relations. Some articles confirmed our results ^(12,13,14,15)^. However, some researchers found the same result, which says traditional risk factors do not correlate to CAV but demonstrate the correlation of hyperlipidemia ^(14)^. Conversely, some results show a correlation between these traditional risk factors, such as hypertension, diabetes, and obesity, and CAV, which is against our results^(16,17,18,19, 23, 37)^.

Although we did not find any relation between smoking and the development of CAV (p-value=0.27), We found smoking can progress CAV in heart transplant patients and increase the probability of PCI in cardiac allograft vasculopathy patients. (p-value= 0.02). Smoking is a significant risk factor for CAV severity. Studies have indicated that smoking might speed up the onset of CAV by inducing endothelial dysfunction, raising oxidative stress levels, and creating an inflammatory environment. These sentinel results verify that exposure to tobacco smoke in both donors and recipients causes vascular inflammation, increased allograft rejection, and graft loss. One of the molecular pathways that overlap and play a role in this phenomenon is the activation of alloimmune cells linked to inflammation caused by tobacco smoke ^(35)^.

Many studies have been performed on the effect of viruses on CAV in heart transplant patients. Results of our research showed a very high association between infectious mononucleosis and CAV in heart transplant patients but not others. Some scientists found an increased expression of antiapoptotic proteins in endothelial cells infected with Epstein-Barr virus (EBV). They hypothesize that the antiapoptotic protein protects against transplant arteriosclerosis. However, the protective effect may be tempered by an increased expression of adhesion molecules, cytokines, and chemokines ^(20,41)^. This hypothesis may be the cause of our results that show the correlation between infectious mononucleosis and CAV.

EBV is a human herpesvirus that causes B cells to become immortal and is linked to the development of many lymphomas, such as DLBCL, HL, and Burkitt’s lymphoma (BL). EBV infection of B cells results in an upregulation of miR-155 expression ^(44,42)^. Loss of miR-155 expression in an EBV-positive DLBCL cell line and in vitro EBV-transformed B cell lines (lymphoblastoid cell lines [LCLs]) decreases cell proliferation and triggers death, suggesting that miR-155 expression is critical for transformed B cell survival ^(45,42)^. In LCLs, miR-155 expression seems to reduce elevated NF-κB activation, which could aid in B cell proliferation and apoptosis prevention ^(46,42)^.

According to some research, there is a high frequency of B lymphocytes infiltrating the coronary arteries of allografts with CAV, which suggests that this process is one of the pathophysiological causes of cardiac allograft vasculopathy ^(43)^.

This could be the reason for our findings that patients with IM had an 8.91-fold increased risk of CAV when compared to other patients. Our research found no correlation between other viruses, such as influenza, Varicella, and herpes viruses, especially CMV, with CAV in heart transplant patients, and some articles confirmed our results ^(21,22,23,24,38)^. Some research demonstrated a correlation between this CMV virus and CAV stimulation of immune response in the vessel wall via induced expression of endothelial MHC antigens and direct endothelial damage resulting in activation of the coagulation cascade, cytokine secretion, and vascular remodeling with vessel lumen loss ^(25,26,27,28,29,30,31,32)^.

However, clinical studies on the association between CMV and CAV have yielded conflicting results. Some studies found no association between recipient age and CAV ^(33)^ nor between recipient gender and CAV ^(14)^. But in our research, we found younger age in Receipt can cause more CAV (P-value= 0.007) and CABG (p-value<0.001) and less mortality (p-value< 0.001), and some studies confirmed it ^(39,40)^. Also, we did not find any gender or race-related CAV in contrast to some smaller studies ^(34–36)^.

## Conclusion

Our study demonstrated that the younger and infectious mononucleosis were significantly associated with developing CAV. Traditional risk factors such as gender, race, hypertension, hyperlipidemia, and diabetes mellitus showed no significant correlation. The older group of recipients has a higher mortality rate compared to the younger one; on the other side, younger age recipients are more likely to do CABG because of CAV. Smoking was associated with a greater need for PCI in CAV patients, indicating that smoking is a contributing factor to the progression of the illness rather than its cause. Association of infectious mononucleosis with CAV was very strong suggesting that EBV may play important role in the pathogenesis of CAV warranting further investigation.

## Funding sources

## Relevant disclosures

## Data Availability

NIS data is publicly available

## Notes

### Competing Interest Statement

The authors have declared no competing interest.

### Clinical Trial

n/a

### Funding Statement

None

### Author Declarations

NIS data base is publicly available without a patient identifier exempt from IRB

## References

(1) Lund, L. H., Edwards, L. B., Dipchand, A. I., Goldfarb, S., Kucheryavaya, A. Y., Levvey, B. J., Meiser, B., Rossano, J. W., Yusen, R. D., & Stehlik, J. (2016b). The Registry of the International Society for Heart and Lung Transplantation: Thirty-third Adult Heart Transplantation Report—2016; Focus Theme: Primary Diagnostic Indications for Transplant. The œJournal of Heart and Lung Transplantation/ the œJournal of Heart and Lung Transplantation, 35(10), 1158–1169. 10.1016/j.healun.2016.08.017

(2) Lee, F., Nair, V., & Chih, S. (2020). Cardiac allograft vasculopathy: Insights on pathogenesis and therapy. Clinical Transplantation/Clinical Transplantation., 34(3). 10.1111/ctr.13794

(3) Bhama, J. K., Nguyen, D. Q., Scolieri, S., Teuteberg, J. J., Toyoda, Y., Kormos, R. L., McCurry, K. R., McNamara, D., & Bermudez, C. A. (2009). Surgical revascularization for cardiac allograft vasculopathy: Is it still an option? Journal of Thoracic and Cardiovascular Surgery/^ the ‰Journal of Thoracic and Cardiovascular Surgery/ the œJournal of Thoracic and Cardiovascular Surgery, 137(6), 1488–1492. 10.1016/j.jtcvs.2009.02.026

(4) Prada-Delgado, O., Estévez-Loureiro, R., López-Sainz, A., Gargallo-Fernández, P., Paniagua-Martín, M., Marzoa-Rivas, R., Barge-Caballero, E., Cuenca-Castillo, J., Castro-Beiras, A., & Crespo-Leiro, M. (2012). Percutaneous coronary Interventions and bypass surgery in patients with cardiac allograft vasculopathy: a Single-Center Experience. Transplantation Proceedings, 44(9), 2657–2659. 10.1016/j.transproceed.2012.09.043

(5) Radovancevic, B., McGiffin, D. C., Kobashigawa, J. A., Cintron, G. B., Mullen, G. M., Pitts, D. E., O’Donnell, J., Thomas, C., Bourge, R. C., & Naftel, D. C. (2003). Retransplantation in 7,290 primary transplant patients: a 10-year multi-institutional study. The œJournal of Heart and Lung Transplantation/ the œJournal of Heart and Lung Transplantation, 22(8), 862–868. 10.1016/s1053-2498(02)00803-3

(6) Luc, J. G. Y., Choi, J. H., Rizvi, S. A., Phan, K., Escrivà, E. M., Patel, S., Reeves, G. R., Boyle, A. J., Entwistle, J. W., Morris, R. J., Massey, H. T., & Tchantchaleishvili, V. (2018). Percutaneous coronary intervention versus coronary artery bypass grafting in heart transplant recipients with coronary allograft vasculopathy: a systematic review and meta-analysis of 1,520 patients. Annals of Cardiothoracic Surgery, 7(1), 19–30. 10.21037/acs.2018.01.10

(7) Kobashigawa, J. A., & Patel, J. K. (2006). Immunosuppression for heart transplantation: where are we now? Nature Clinical Practice Cardiovascular Medicine, 3(4), 203–212. 10.1038/ncpcardio0510

(8) Taylor, D. O., Stehlik, J., Edwards, L. B., Aurora, P., Christie, J. D., Dobbels, F., Kirk, R., Kucheryavaya, A. Y., Rahmel, A. O., & Hertz, M. I. (2009). Registry of the International Society for Heart and Lung Transplantation: Twenty-sixth Official Adult Heart Transplant Report—2009. the œJournal of Heart and Lung Transplantation/ the œJournal of Heart and Lung Transplantation, 28(10), 1007–1022. 10.1016/j.healun.2009.08.014

(9) Costanzo, M. R., Dipchand, A., Starling, R., Anderson, A., Chan, M., Desai, S., Fedson, S., Fisher, P., Gonzales-Stawinski, G., Martinelli, L., McGiffin, D., Smith, J., Taylor, D., Meiser, B., Webber, S., Baran, D., Carboni, M., Dengler, T., Feldman, D., . . . Vanhaecke, J. (2010). The International Society of Heart and Lung Transplantation Guidelines for the care of heart transplant recipients. The œJournal of Heart and Lung Transplantation/ the œJournal of Heart and Lung Transplantation, 29(8), 914–956. 10.1016/j.healun.2010.05.034

(10) Libby, P., Lichtman, A.H. and Hansson, G.K., 2013. Immune effector mechanisms implicated in atherosclerosis: from mice to humans. Immunity, 38(6), pp.1092–1104.

(11) Kaczmarek, I., Deutsch, M.A., Rohrer, M.E., Beiras-Fernandez, A., Groetzner, J., Daebritz, S., Schmoeckel, M., Spannagl, M., Meiser, B. and Reichart, B., 2006. HLA-DR matching improves survival after heart transplantation: is it time to change allocation policies?. The Journal of heart and lung transplantation, 25(9), pp.1057–1062.

(12) Pober, J. S., Jane-Wit, D., Qin, L., & Tellides, G. (2014). Interacting mechanisms in the pathogenesis of cardiac allograft vasculopathy.(8), 1609–

(13) Fluschnik, N., Geelhoed, B., Becher, P. M., Schrage, B., Brunner, F. J., Knappe, D., Bernhardt, A. M., Blankenberg, S., Kobashigawa, J., Reichenspurner, H., Schnabel, R. B., & Magnussen, C. (2021). Non-immune risk predictors of cardiac allograft vasculopathy: Results from the U.S. organ procurement and transplantation network. International Journal of Cardiology, 331, 57–62. 10.1016/j.ijcard.2021.02.002

(14) Lázaro, I., Bonet, L. A., López, J. M., Lacuesta, E., Martínez-Dolz, L., Ramón-Llín, J. A., Lalaguna, L. A., Pérez, O. C., Martínez, V. O., Fuentes, F. B., & Sanz, A. S. (2008). Influence of traditional cardiovascular risk factors in the recipient on the development of cardiac allograft vasculopathy after heart transplantation. Transplantation Proceedings, 40(9), 3056–3057. 10.1016/j.transproceed.2008.08.115

(15) Chang, G., DeNofrio, D., Desai, S., Kelley, M. P., Rader, D. J., Acker, M. A., & Loh, E. (1998). Lipoprotein(a) levels and heart transplantation atherosclerosis. American Heart Journal, 136(2), 329–334. 10.1053/hj.1998.v136.89581

(16) Raichlin, E. R., McConnell, J. P., Lerman, A., Kremers, W. K., Edwards, B. S., Kushwaha, S. S., Clavell, A. L., Rodeheffer, R. J., & Frantz, R. P. (2007). Systemic inflammation and metabolic syndrome in cardiac allograft vasculopathy. The Journal of Heart and Lung Transplantation, 26(8), 826–833. 10.1016/j.healun.2007.05.008

(17) Sánchez-Gómez, J. M., Martínez-Dolz, L., Sánchez-Lázaro, I., Almenar, L., Sánchez- Lacuesta, E., Muñoz-Giner, B., Portolés, M., Rivera, M., Valera-Román, A., González-Juanatey, J. R., Tejada-Ponce, D., Agüero, J., Buendía, F., & Salvador, A. (2011). Influence of metabolic syndrome on development of cardiac allograft vasculopathy in the transplanted heart. Transplantation, 93(1), 106–111. 10.1097/tp.0b013e3182398058

(18) Kato, T., Chan, M. C., Gao, S., Schroeder, J. S., Yokota, M., Murohara, T., Iwase, M., Noda, A., Hunt, S. A., & Valantine, H. A. (2004). Glucose intolerance, as reflected by hemoglobin a1clevel, is associated with the incidence and severity of transplant coronary artery disease. Journal of the American College of Cardiology, 43(6), 1034–1041. 10.1016/j.jacc.2003.08.063

(19) Valantine, H., Rickenbacker, P., Kemna, M., Hunt, S., Chen, Y. I., Reaven, G., & Stinson, E. B. (2001). Metabolic abnormalities characteristic of dysmetabolic syndrome predict the development of transplant coronary artery disease. Circulation, 103(17), 2144–2152. 10.1161/01.cir.103.17.2144

(20) Gatto, G., Rossi, A., Rossi, D., Kroening, S., Bonatti, S., & Mallardo, M. (2008). Epstein– Barr virus latent membrane protein 1 trans-activates miR-155 transcription through the NF-κB pathway. Nucleic Acids Research, 36(20), 6608–6619. 10.1093/nar/gkn666

(21) Potena, L., Masetti, M., Russo, A., & Grigioni, F. (2016). Current perspectives on cytomegalovirus in heart transplantation. Current Transplantation Reports, 3(4), 358–366. 10.1007/s40472-016-0121-x

(22) Luckraz, H., Charman, S. C., Wreghitt, T., Wallwork, J., Parameshwar, J., & Large, S. R. (2003). Does cytomegalovirus status influence acute and chronic rejection in heart transplantation during the ganciclovir prophylaxis era? The Journal of Heart and Lung Transplantation, 22(9), 1023–1027. 10.1016/s1053-2498(02)01185-3

(23) Pober, J. S., Chih, S., Kobashigawa, J., Madsen, J. C., & Tellides, G. (2021, November 22). Cardiac allograft vasculopathy: Current review and future research directions. Cardiovascular research. https://www.ncbi.nlm.nih.gov/pmc/articles/PMC8783389/

(24) Srivastava, R., Curtis, M., Hendrickson, S., Burns, W. H., & Hosenpud, J. D. (1999). STRAIN SPECIFIC EFFECTS OF CYTOMEGALOVIRUS ON ENDOTHELIAL CELLS: IMPLICATIONS FOR INVESTIGATING THE RELATIONSHIP BETWEEN CMV AND CARDIAC ALLOGRAFT VASCULOPATHY1. Transplantation, 68(10), 1568–1573. 10.1097/00007890-199911270-00022

25. (25) Petrakopoulou, P., KüBrich, M., Pehlivanli, S., Meiser, B., Reichart, B., Von Scheidt, W., & Weis, M. (2004). Cytomegalovirus infection in heart transplant recipients is associated with impaired endothelial function. Circulation, 110(11_suppl_1). 10.1161/01.cir.0000138393.99310.1c

(26) Weill, D. (2001). Role of cytomegalovirus in cardiac allograft vasculopathy. Transplant Infectious Disease, 3(s2), 44–48. 10.1034/j.1399-3062.2001.00009.x

(27) Schmauss, D., & Weis, M. (2008). Cardiac allograft vasculopathy. Circulation, 117(16), 2131–2141. 10.1161/circulationaha.107.711911

(28) Sharples, L. D., Jackson, C. H., Parameshwar, J., Wallwork, J., & Large, S. R. (2003). Diagnostic accuracy of coronary angiography and risk factors for post–heart-transplant cardiac allograft vasculopathy. Transplantation, 76(4), 679–682. 10.1097/01.tp.0000071200.37399.1d

29. (29) Weis, M., & Von Scheidt, W. (1997). Cardiac allograft vasculopathy. Circulation, 96(6), 2069–2077. 10.1161/01.cir.96.6.2069

(30) Fateh-Moghadam, S., Bocksch, W., Wessely, R., Jäger, G., Hetzer, R., & Gawaz, M. (2003). Cytomegalovirus infection status predicts progression of heart-transplant vasculopathy. Transplantation, 76(10), 1470–1474. 10.1097/01.tp.0000090163.48433.48

(31) Bonaros, N. E., Kocher, A., Dunkler, D., Grimm, M., Zuckermann, A., Ankersmit, J., Ehrlich, M., Wolner, E., & Laufer, G. (2004). Comparison of combined prophylaxis of cytomegalovirus hyperimmune globulin plus ganciclovir versus cytomegalovirus hyperimmune globulin alone in high-risk heart transplant recipients1. Transplantation, 77(6), 890–897. 10.1097/01.tp.0000119722.37337.dc

(32) Johansson, I., Andersson, R., Friman, V., Selimovic, N., Hanzen, L., Nasic, S., Nyström, U., & Sigurdardottir, V. (2015). Cytomegalovirus infection and disease reduce 10-year cardiac allograft vasculopathy-free survival in heart transplant recipients. BMC Infectious Diseases, 15(1). 10.1186/s12879-015-1321-1

(33) Gao, S., Hunt, S. A., Alderman, E. L., Liang, D., Yeung, A. C., & Schroeder, J. S. (1997). Relation of donor age and preexisting coronary artery disease on angiography and intracoronary ultrasound to later development of accelerated allograft coronary artery disease. Journal of the American College of Cardiology, 29(3), 623–629. 10.1016/s0735-1097(96)00521-9

(34) Moayedi, Y., Fan, C. P. S., Cherikh, W. S., Stehlik, J., Teuteberg, J. J., Ross, H. J., & Khush, K. K. (2019). Survival outcomes after heart transplantation. Circulation Heart Failure, 12(10). 10.1161/circheartfailure.119.006218

(35) Khanna, A. K., Xu, J., Uber, P. A., Burke, A. P., Baquet, C., & Mehra, M. R. (2009). Tobacco smoke exposure in either the donor or recipient before transplantation accelerates cardiac allograft rejection, vascular inflammation, and graft loss. Circulation, 120(18), 1814–1821. 10.1161/circulationaha.108.840223

(36) Khush, K. K., Cherikh, W. S., Chambers, D. C., Harhay, M. O., Hayes, D., Hsich, E., Meiser, B., Potena, L., Robinson, A., Rossano, J. W., Sadavarte, A., Singh, T. P., Zuckermann, A., & Stehlik, J. (2019). The International Thoracic Organ Transplant Registry of the International Society for Heart and Lung Transplantation: Thirty-sixth adult heart transplantation report — 2019; focus theme: Donor and recipient size match. The Journal of Heart and Lung Transplantation, 38(10), 1056–1066. 10.1016/j.healun.2019.08.004

(37) Erinc, K., Yamani, M. H., Starling, R. C., Crowe, T., Hobbs, R., Bott-Silverman, C., Rincon, G., Young, J. B., Feng, J., Cook, D. J., Smedira, N., & Tuzcu, E. M. (2005). The effect of combined Angiotensin-Converting enzyme inhibition and calcium antagonism on allograft coronary vasculopathy validated by intravascular ultrasound. The Journal of Heart and Lung Transplantation, 24(8), 1033–1038. 10.1016/j.healun.2004.06.005

(38) Dummer, S., Lee, A., Breinig, M. K., Kormos, R., Ho, M., & Griffith, B. (1994). Investigation of cytomegalovirus infection as a risk factor for coronary atherosclerosis in the explanted hearts of patients undergoing heart transplantation. Journal of Medical Virology, 44(3), 305–309. 10.1002/jmv.1890440316

(39) Shetty, M., & Chowdhury, Y. S. (2023, May 22). Heart transplantation allograft vasculopathy. StatPearls - NCBI Bookshelf. https://www.ncbi.nlm.nih.gov/books/NBK555999/

(40) Transplant coronary disease: nonimmunologic risk factors. (1992, June 1). PubMed. https://pubmed.ncbi.nlm.nih.gov/1622991/

(41) Hatton, O., Smith, M. M., Alexander, M., Mandell, M., Sherman, C., Stesney, M. W., Hui, S. T., Dohrn, G., Medrano, J., Ringwalt, K., Harris-Arnold, A., Maloney, E. M., Krams, S. M., & Martinez, O. M. (2019). Epstein-Barr Virus Latent Membrane Protein 1 Regulates Host B Cell MicroRNA-155 and Its Target FOXO3a via PI3K p110α Activation. Frontiers in Microbiology, 10. 10.3389/fmicb.2019.02692

(42) Wood, C. D., Carvell, T., Gunnell, A., Ojeniyi, O. O., Osborne, C., & West, M. J. (2018). Enhancer control of MicroRNA MIR-155 expression in Epstein-Barr Virus-Infected B cells. Journal of Virology, 92(19). 10.1128/jvi.00716-18

(43) Chatterjee, D., Moore, C., Gao, B., Clerkin, K. J., See, S. B., Shaked, D., Rogers, K., Nunez, S., Veras, Y., Addonizio, L., Givertz, M. M., Naka, Y., Mancini, D., Vasilescu, R., Marboe, C., Restaino, S., Madsen, J. C., & Zorn, E. (2017). Prevalence of polyreactive innate clones among graft-infiltrating B cells in human cardiac allograft vasculopathy. The Journal of Heart and Lung Transplantation, 37(3), 385–393. 10.1016/j.healun.2017.09.011

(44) Mrazek, J., Kreutmayer, S. B., Grasser, F. A., Polacek, N., & Huttenhofer, A. (2007). Subtractive hybridization identifies novel differentially expressed ncRNA species in EBV-infected human B cells. Nucleic Acids Research, 35(10), e73. 10.1093/nar/gkm244

(45) Linnstaedt, S. D., Gottwein, E., Skalsky, R. L., Luftig, M. A., & Cullen, B. R. (2010). Virally induced cellular MicroRNA MIR-155 plays a key role in B-Cell immortalization by Epstein-Barr virus. Journal of Virology, 84(22), 11670–11678. 10.1128/jvi.01248-10

(46) Lu, F., Weidmer, A., Liu, C., Volinia, S., Croce, C. M., & Lieberman, P. M. (2008). Epstein-Barr Virus-Induced MIR-155 attenuates NF-ΚB signaling and stabilizes latent virus persistence. Journal of Virology, 82(21), 10436–10443. 10.1128/jvi.00752-08

